# Evaluating a Multitask AI Model versus Humans for Portion Size Estimation

**DOI:** 10.64898/2026.04.16.26351036

**Authors:** Bibinur Nurmanova, Zhuldyz Omarova, Aibota Sanatbyek, Huseyin Atakan Varol, Mei-Yen Chan

**Affiliations:** School of Medicine, Nazarbayev University, Astana 010000, Kazakhstan; Institute of Smart Systems and Artificial Intelligence, Nazarbayev University, Astana 010000, Kazakhstan

**Keywords:** food portion size estimation, dietary assessment, Central Asia, digital visual food atlas, artificial intelligence, image recognition, benchmarking, machine learning, AI-driven nutrition

## Abstract

**Background:** Accurate dietary assessment is essential for precision nutrition and effective nutrition surveillance. However, portion size estimation remains a persistent challenge, particularly in culturally diverse regions such as Central Asia. Traditional self-reporting tools often yield inconsistent results due to communal eating practices and unfamiliarity with standard measures.

**Objective:** To address these limitations, this study aimed to compare three methods: unassisted human judgment, visual food atlas assistance, and an artificial intelligence (AI) model, using Central Asian food items.

**Methods:** In this cross-sectional study, 128 participants from Astana, Kazakhstan, visually estimated portion sizes of 51 foods and 8 beverages from standardized photographs. Participants were randomized into two groups: one using unassisted visual estimation and the other aided by a regionally tailored digital food atlas. Additionally, an AI model trained on Central Asian food images was evaluated. Actual food weights served as the reference standard. Accuracy was assessed using Mean Absolute Error (MAE) and Mean Absolute Percentage Error (MAPE) across food types and portion sizes.

**Results:** The atlas-assisted group demonstrated the highest accuracy, with the lowest MAE (80.81g) and MAPE (44.76%) across all portions. The AI model showed promising results for average portions (MAE: 79.07g, MAPE: 67.91%) but underperformed on small portions, particularly for meat-based items. Unassisted estimates were the least accurate (MAE: 133.86g, MAPE: 79.40%). Across food categories, visual aids consistently improved accuracy, while AI demonstrated variability by texture and portion size.

**Conclusions:** Culturally adapted visual atlases significantly enhance portion size estimation accuracy in non-Western, communal-eating contexts. While AI models hold promise for dietary assessments, particularly with standard portions and beverages, further refinement is needed for complex food items and small portion types. These findings support the integration of visual and AI-based tools into region-specific dietary monitoring strategies.

## Introduction

Dietary habits in Central Asia are undergoing a complex, multifaceted transition shaped by the convergence of modernization and deep-rooted cultural traditions (1,2). In the urban areas of Azerbaijan, Kazakhstan, Kyrgyzstan, Tajikistan, Turkmenistan, and Uzbekistan, the consumption of calorie-dense, ultra-processed foods, high in sugar, fat, and salt, is increasingly prevalent, further contributing to significant dietary risks in these regions (3–5). On the other hand, rural populations continue to rely more on long-standing cultural dietary practices (6), which often favor higher consumption of red meat, meat products, and certain grains with a lower intake of fruits and vegetables (7–11).

The perpetuation of these culturally ingrained food habits, which have focused on long-standing traditions rather than being guided by modern evidence-based dietary guidance, has led to a dual burden of malnutrition. This includes a rising prevalence of non-communicable diseases (NCDs), such as obesity, hypertension, and type 2 diabetes, coexisting with micronutrient deficiencies in some areas of these countries (7–9,12–15). Recent work has also highlighted the need for region-specific anthropometric cut-offs, such as body mass index (BMI) and waist circumference, tailored to Central Asian populations (16). Despite this growing burden of diet-related risk factors, efforts to change dietary habits remain hindered by a lack of accurate, robust dietary data. The main problem lies in the inadequacy of current dietary assessment tools. Most existing instruments were developed in and for Western contexts, with little adaptation to communal eating habits, non-standard portioning, or food items commonly consumed in Central Asia (3–5,7,10,17). As a result, this misalignment leads to inaccurate estimates of nutrient intake, undermines the development of evidence-based policies, and limits the effectiveness of nutrition and dietary interventions in public healthcare settings. This challenge is compounded by limited nutrition competencies among healthcare professionals in Kazakhstan, with recent evidence showing gaps between knowledge and its clinical application (18).

In response, researchers have explored two technological advances: artificial intelligence (AI) - based image recognition systems and visual food atlases. Over the past decade, numerous studies have examined AI in dietary assessment, particularly for portion size estimation. AI-based tools, including image recognition models, augmented reality (AR) interfaces, and immersive digital dietitians, have shown promising results in improving accuracy and reducing respondent burden. For example, Whitton et al. (19) found that AI-based or image-assisted systems offer a more precise and less burdensome solution for dietary surveillance and reduce overestimation biases common in interviewer-assisted methods. Similarly, Braga et al. (20) showed that the immersive virtual alimentation and nutrition (IVAN) dietitian, delivered through immersive virtual reality (iVR), significantly improved participants’ portion size estimation accuracy and self-efficacy, particularly for high-energy-dense foods. While the impact was comparable to in-person education, the scalability and engagement levels offered by the digital tool present a major advantage in diverse or resource-limited contexts. A broader review of AI-based dietary assessment tools indicates that image-based deep learning models outperform traditional methods when trained on context-specific data. However, most systems have only been validated in Western settings, assuming individual portioning and familiarity with grams or cups. These models have not been sufficiently tested in communal-eating populations, such as those in Central Asia, where meals are served on large platters and shared informally without clearly defined personal portions (21). As a result, their real-world applicability and accuracy remain uncertain.

Regionally adapted visual food atlases have emerged as complementary strategies to address these cultural and contextual gaps. When modified to include local foods, typical serving ware, and realistic portion sizes, food atlases have improved estimation accuracy in non-Western settings, including the Middle East, North Africa, and the Balkans (22,23). For Central Asian populations, where nutritional literacy may be limited and standardized portioning uncommon, a visual reference tailored to local food habits could offer a practical solution. Studies show that communal or buffet-style eating, especially self-served or shared-plate formats, significantly increases the error and variability of portion-size and nutrient intake estimation compared to individually plated meals, with systematic underestimation or wide random error documented in children and adults (24,25). In such contexts, a digital food atlas designed around region-specific foods and eating practices may help reduce misestimation and improve dietary assessment accuracy.

Despite their usefulness, food atlases rely on human interpretation and are difficult to scale. In contrast, AI-based image recognition systems can offer automated, real-time portion estimation with minimal user input and are highly scalable. Both tools are especially relevant in Central Asia, where dietary cultures are shared across borders, but access to trained dietitians and validated assessment tools remains limited. While each method has advantages, it remains unclear which performs better in real-world, culturally specific, low-resource, or communal-eating settings. Most prior research compares AI to recall or interviewer-based methods (e.g., Whitton et al., 2024), rather than specifically to visual atlases. Therefore, a key knowledge gap remains on whether AI can offer a better alternative to visual estimation via food atlases, especially in regions like Central Asia, where scalable solutions are urgently needed.

This study aimed to evaluate and compare three portion size estimation methods: (1) unassisted human judgment, (2) a newly developed Central Asia–specific digital visual food atlas, and (3) an AI model trained on over 21,000 regionally sourced food images. Our contributions included: validating the Central Asia-specific visual food atlas with ground-truth weight measurements of food items and beverages; assessing the AI model’s accuracy and providing future performance recommendations; and developing a standardized dietary assessment tool for Central Asia to support researchers, dietitians, and healthcare professionals.

## Materials and Methods

### Study Design

This cross-sectional comparative validation study assessed the accuracy of three portion size estimation methods: unassisted visual estimation, a culturally adapted visual food atlas, and an AI-based image analysis model.

### Study Population

A total of 128 adult participants aged 18 to 70 years were recruited from Astana, Kazakhstan, through convenience sampling via a public call. Individuals with professional training in food estimation, such as dietitians, nutritionists, and chefs, were excluded (unless they were part of the atlas-assisted group) to ensure that unassisted estimates reflected the general population’s ability.

The sample size was calculated using the following parameters: a detectable difference (d) of 5%, a standard deviation (σ) of 10, a significance level (α) = 0.05 (Zα/2 = 1.96), and power = 0.80 (Zβ = 0.84) (26,27). Based on previous studies (28–31), a relative difference of 5% between estimated and actual serving sizes was considered meaningful. After accounting for a 25% dropout rate (19,29,32), the required sample size per group was calculated using the following formula (33):

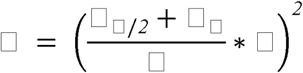

A minimum of 32 participants per group, or 64 participants overall, were needed. To ensure this number of responses by the end of this study, at least 43 participants per group were recruited.

### Comparison Groups

Participants were randomly assigned to one of two human estimation groups:

1. **Unassisted group.** Participants estimated food portion sizes without any visual aids. They relied solely on their perceptions when answering questions about food volumes. This group served as a baseline for comparison against the atlas-assisted group and the AI model.
2. **Atlas-assisted group.** Participants received a culturally adapted visual food atlas that included photographs of three portion sizes for each food item. The shortened version of the food atlas provided to participants featured 60 food types and 8 beverages (totaling 68 items). The full version included 115 common food items, consisting of 95 photo series and 20 guide photographs of beverages. The selection of food categories and items was based on the Central Asian Food Dataset (CAFD) and Central Asian Food Scenes Dataset (CAFSD), which offered a comprehensive collection of over 16,000 and 21,306 images, respectively (34,35). A previous study (32) found no significant differences between estimations using digital and printed photo series. Therefore, the food atlas was integrated digitally into the online questionnaire. Example pages of the food atlas are shown in **Figure 1**.

**Figure 1:**
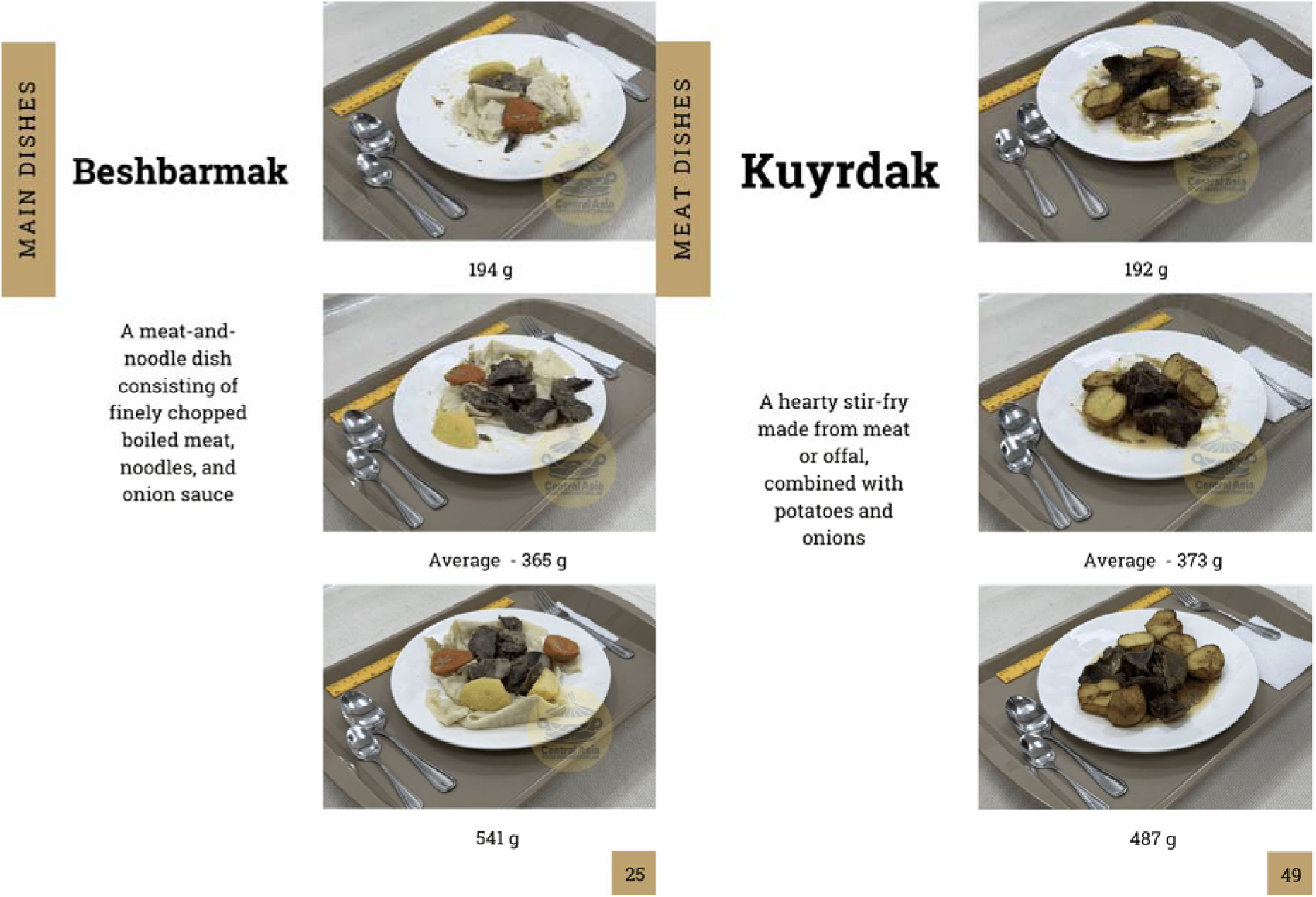
This figure shows representative screenshots from the shortened Central Asian visual food atlas used in the atlas-assisted group. Each series displayed three portion sizes (small, medium, large) of locally consumed foods. Participants used these standardized images to estimate portion sizes.

Additionally, a third comparison group was established based on:

3. **AI model.** The AI model was trained for the specific task of food volume estimation using an image dataset of common Central Asian food items paired with corresponding ground-truth volumes. A YOLOv12-based end-to-end multi-task framework was applied, integrating both food detection and portion size estimation within a single architecture. The model leveraged pre-trained weights from the recently introduced Food Portion Benchmark (FPB) dataset (36), which consists of 14,083 images across 138 food classes with laboratory-measured component weights. These weights were fine-tuned on Central Asian food categories derived from the CAFD and CAFSD, ensuring contextual and cultural relevance. This approach enabled the model to achieve accurate portion estimation for diverse regional foods while maintaining adaptability to the complexities of communal eating practices.

### Data Collection

The survey included photographs of 51 food types and 8 beverages, presented in three portion sizes (small, medium, and large). This set of representative food items was selected to reflect a broad range of appearances, consistencies, and textures (29). This approach ensured adequate coverage of different food categories while also including a small number of items outside the original atlas to minimize recognition bias and enhance the validity of portion size estimation.

The survey was deployed via the Qualtrics platform (Qualtrics International Inc., March 2025 version). Each participant received a personalized survey link with a randomized selection of 9 food items and 3 beverages, ensuring varied exposure while minimizing participant fatigue. Each portion was evaluated by at least three different participants. Potential sources of bias included convenience sampling and familiarity with certain foods. Randomization of food items was used to minimize ordering bias.

Participants viewed standardized food images and used a slider to estimate portion sizes in grams or milliliters (**Figure 2**). Slider ranges were based on the minimum and maximum actual weights or volumes of the items. Participants also reported their confidence level (low, medium, or high) and left optional comments on any difficulties or uncertainties.

**Figure 2:**
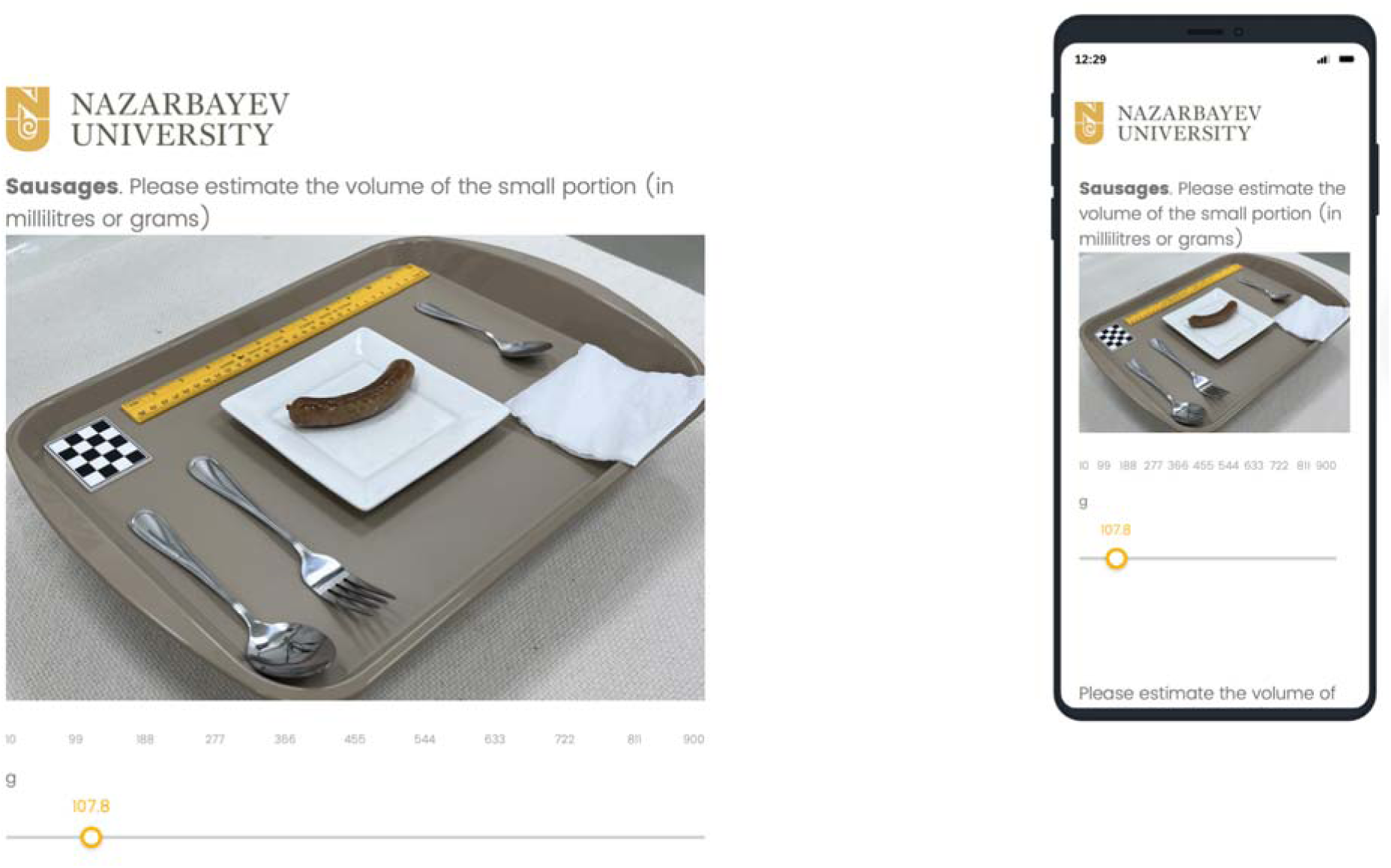
This figure is an example of the online survey platform (Qualtrics International Inc.) showing the slider tool used by participants to provide gram or milliliter estimates of portion sizes. Slider ranges were based on actual minimum and maximum measured values.

In the AI assessment, the same food images were analyzed by the AI model, which generated estimated weights based on its training data and model architecture.

### Data Analysis

Descriptive statistics (frequencies, percentages, means, and standard deviations) were used to summarize demographic data. Ground-truth food weights were recorded and used as reference values for validation.

Estimation accuracy was assessed using two primary error metrics, such as Mean Absolute Error (MAE) and Mean Absolute Percentage Error (MAPE). Absolute differences in grams were evaluated with MAE to show the magnitude of the typical error and report overall deviation regardless of under- or overestimation. MAPE was used for relative error assessment and cross-comparison of food items of different sizes and weights, which is helpful when dealing with a variety of foods.

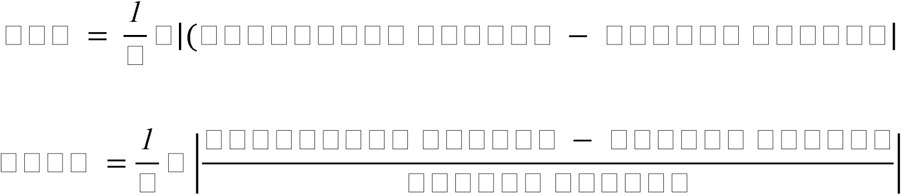

The primary outcome was estimation error (MAE, MAPE). These metrics allowed the comparison of human and AI estimates with actual portion sizes. No confounders or effect modifiers were analyzed. Results were stratified by group, food category, and portion size.

### Ethical Considerations

Ethical approval (2024Sep#01) was obtained from the Nazarbayev University School of Medicine Institutional Research Ethics Committee (NUSOM IREC). All research design, practices, and reporting of studies were conducted in alignment with the NUSOM IREC Bylaws, NUSOM IREC Procedures, and other NU institutional policies regarding research ethics.

## Results

### Characteristics of the study population

A total of 128 participants completed the study and were included in the final analysis. Gender and age distributions for the unassisted and atlas-assisted groups are presented in **Table 1**.

**Table 1.**
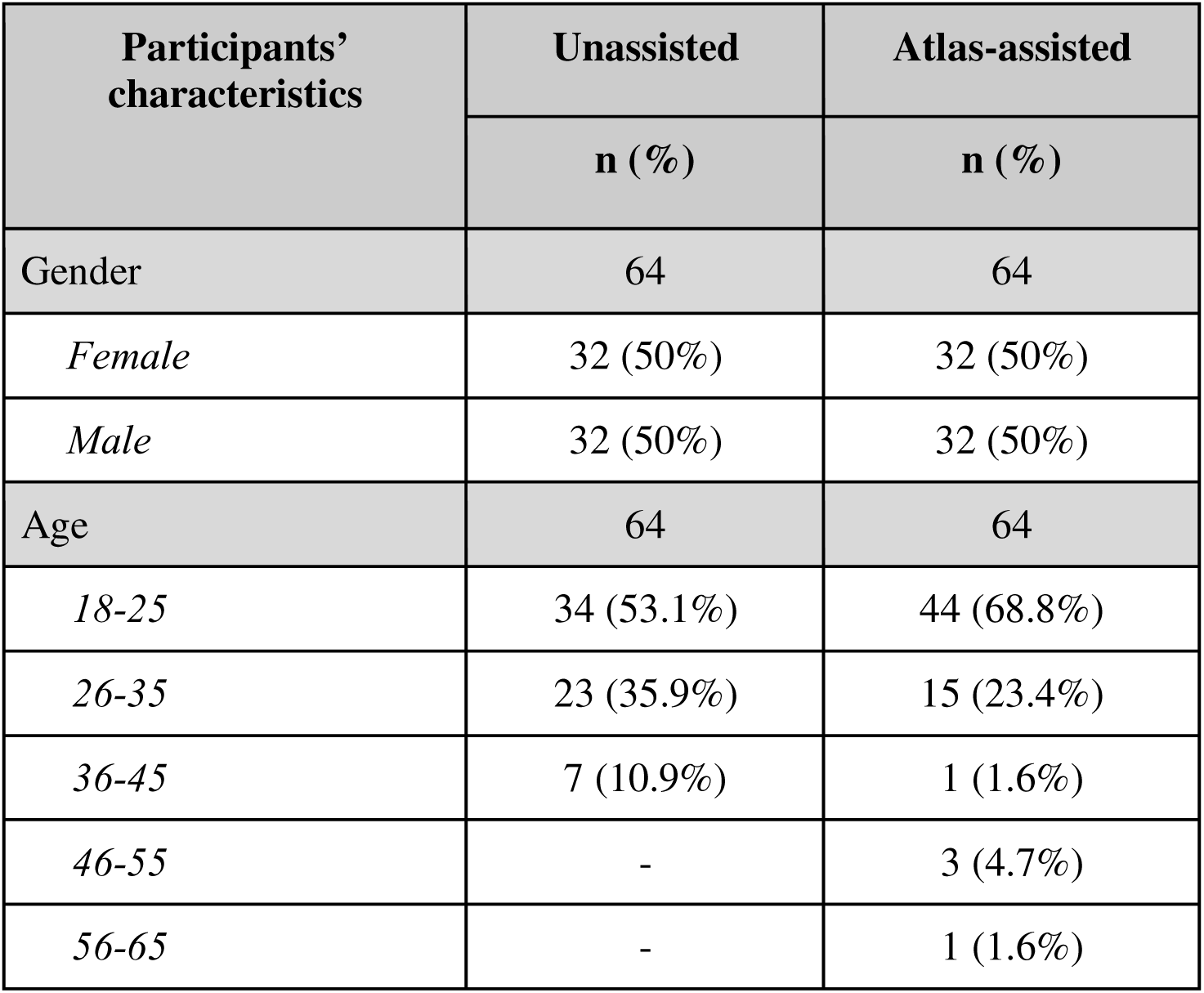
Demographic characteristics of human evaluators.

| Participants' characteristics | Unassisted | Atlas-assisted |
| --- | --- | --- |
|  | n (%) | n (%) |
| Gender | 64 | 64 |
| *Female* | 32 (50%) | 32 (50%) |
| *Male* | 32 (50%) | 32 (50%) |
| Age | 64 | 64 |
| *18-25* | 34 (53.1%) | 44 (68.8%) |
| *26-35* | 23 (35.9%) | 15 (23.4%) |
| *36-45* | 7 (10.9%) | 1 (1.6%) |
| *46-55* | - | 3 (4.7%) |
| *56-65* | - | 1 (1.6%) |

### Comparison of Estimation Methods by Error Metrics

**Table 2** summarizes the accuracy of the unassisted, atlas-assisted, and AI Model methods based on mean, standard deviation (SD), MAE, and MAPE, stratified by portion size. “Standard portion” referred to the average portion of beverages.

**Table 2.** MAE and MAPE results obtained from unassisted, atlas-assisted, and AI model methods.

|  | Unassisted |  |  | Atlas-assisted |  |  | AI Model |  |
| --- | --- | --- | --- | --- | --- | --- | --- | --- |
| | Mean ±<br>SD (g) | MAE<br>(g) | MAPE<br>(%) | Mean ±<br>SD (g) | MAE<br>(g) | MAPE<br>(%) | MAE<br>(g) | MAPE<br>(%) |
| **Overall** | 249.75 ±<br>187.98 | 133.86 | 79.40 | 214.81 ±<br>133.75 | 80.81 | 44.76 | 97.35 | 85.71 |
| **Small**<br>**portion** | 152.63 ±<br>64.93 | 86.65 | 94.30 | 120.29<br>± 42.66 | 45.82 | 47.94 | 102.06 | 151.50 |
| **Average**<br>**portion** | 252.40 ±<br>78.76 | 140.69 | 82.21 | 208.53<br>± 67.12 | 83.02 | 46.87 | 79.07 | 67.91 |
| **Large**<br>**portion** | 357.23 ±<br>93.10 | 187.73 | 69.73 | 311.51<br>± 94.23 | 119.18 | 42.66 | 116.23 | 46.88 |
| **Standard**<br>**portion** | 214.93 ±<br>22.76 | 84.14 | 35.39 | 227.24<br>± 43.44 | 39.12 | 16.89 | 56.51 | 16.94 |

As shown, the atlas-assisted method demonstrated the highest accuracy, achieving the lowest MAE and MAPE across most portion sizes. The AI model performed comparably for average and large portions but showed a marked drop in accuracy for small portions. The unassisted group showed the least accurate estimations overall.

### Food-Specific Estimation Errors

**Figure 3** presents the MAE in grams across individual food items across the unassisted, atlas-assisted, and AI model methods.

**Figure 3:**
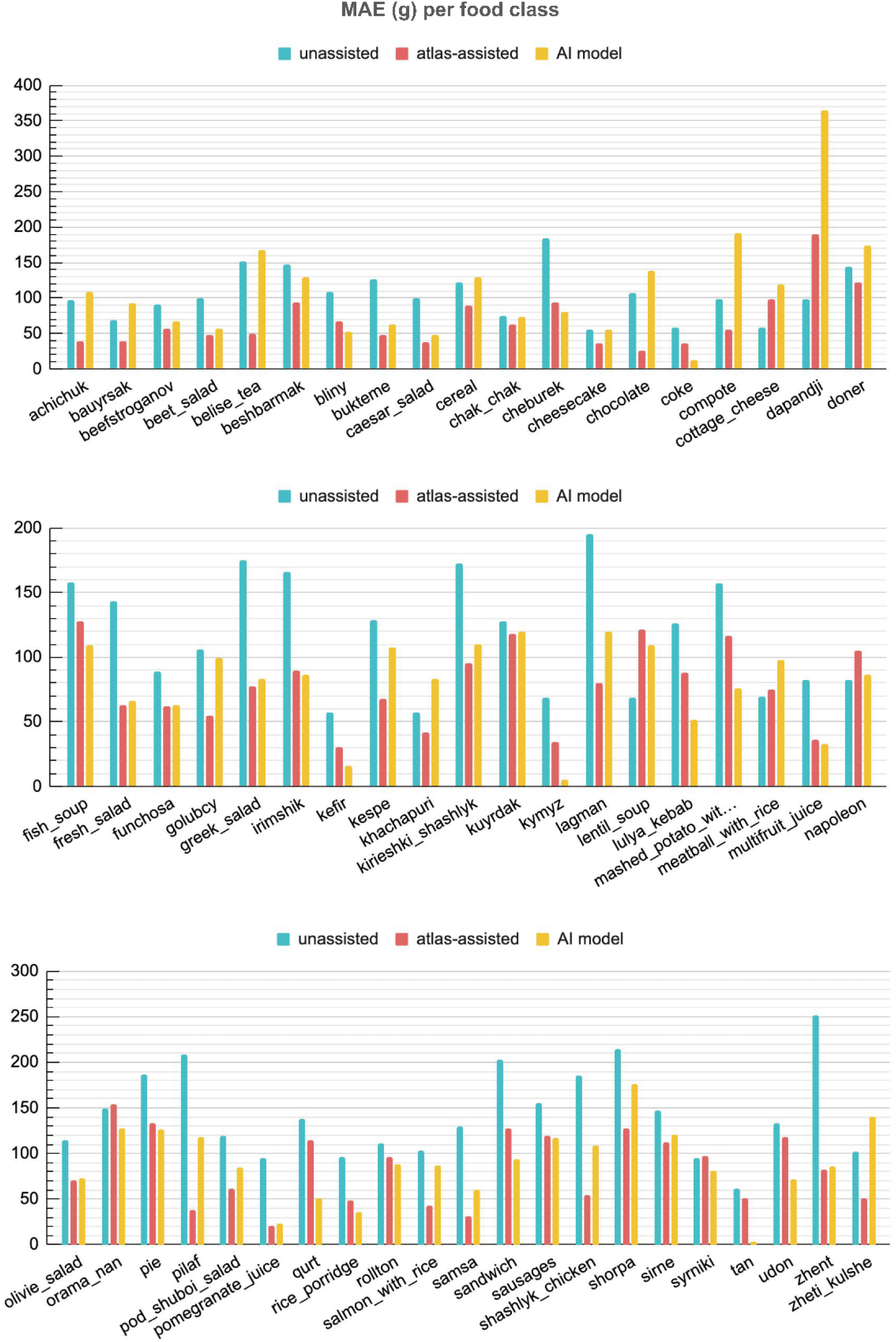
This figure shows the mean absolute error (MAE) of portion size estimation across individual food items. Comparison of MAE values (in grams) was done for the unassisted, atlas-assisted, and AI model groups.

Across most items, atlas-assisted estimation resulted in lower MAE values compared with unassisted estimation, indicating improved accuracy when culturally tailored visual references were provided. This was particularly evident for structured or commonly consumed dishes such as *shashlyk_chicken*, *pilaf*, and *samsa*, where visual cues likely aided recognition of standard serving quantities.

The AI-assisted method demonstrated variable performance. For some foods, especially dense or single-component items such as *qurt*, *cheburek*, and *lulya_kebab,* the AI model achieved lower MAE values than both unassisted and atlas-assisted estimates. Additionally, the AI model performed well with beverages, as observed in items such as *coke, kymyz, kefir,* and *tan.* However, for other items, particularly amorphous foods (those without a fixed shape or structure), salads, or liquid-based dishes, such as soups, AI-derived estimates were less accurate, often exceeding the MAE achieved by atlas-assisted estimation. This pattern may have reflected limitations in the model’s ability to distinguish volume and texture variations from two-dimensional images.

Unassisted estimation consistently produced the highest MAE values across most food classes. Notably, MAEs exceeded 150 g for items such as *lagman*, *greek_salad*, *zhent*, and *irimshik*, highlighting the difficulty of estimating portion size without visual references in a region where standardized servings are uncommon and foods are often shared.

In summary, the visual atlas-assisted method provided the most consistently accurate portion estimates across food types, particularly for composite and familiar dishes. The AI model showed comparable or superior performance for some items, especially dense or easily segmentable foods and beverages, but performance was inconsistent for less structured or volume-sensitive dishes. These findings suggest that while AI-assisted tools hold promise, further refinement and food-specific calibration may be necessary before widespread implementation in communal-eating contexts such as Central Asia.

### Category-Wise Accuracy Comparison

**Figure 4** presents the MAE and MAPE for each estimation method across nine food categories. Food items were grouped into categories:

● Meat-based: *beefstroganov, mashed_potato_with_chicken_in_plum, dapanji, lulya_kebab, meatball_with_rice, pilaf, sausage, shashlyk_chicken, sirne;*
● Dairy-based: *beshbarmak, cottage cheese, irimshik, kefir, kurt, kymyz, tan, zhent*;
● Flour-based: *bauyrsak, bliny, bukteme, cheburek, khachapuri, samsa, zheti_kulshe;*
● Desserts: *chak_chak, cheesecake, napoleon, kirieshki_shashlyk, pie, chocolate, syrniki;*
● Salads: *olivie_salad, greek_salad, caesar_salad, funchosa, pod_shuboi_salad, beet_salad, fresh_salad, achichuk;*
● Mixed dishes: *doner, kespe, rollton, fish_soup, lentil_soup, salmon_with_rice, sandwich, shorpa, udon;*
● Main dishes: *lagman, orama_nan, kuyrdak, golubcy;*
● Drinks: *belise_tea, multifruit_juice, coke, compote, pomegranate_juice;*
● Cereals: *rice_porridge, cereals*.

**Figure 4:**
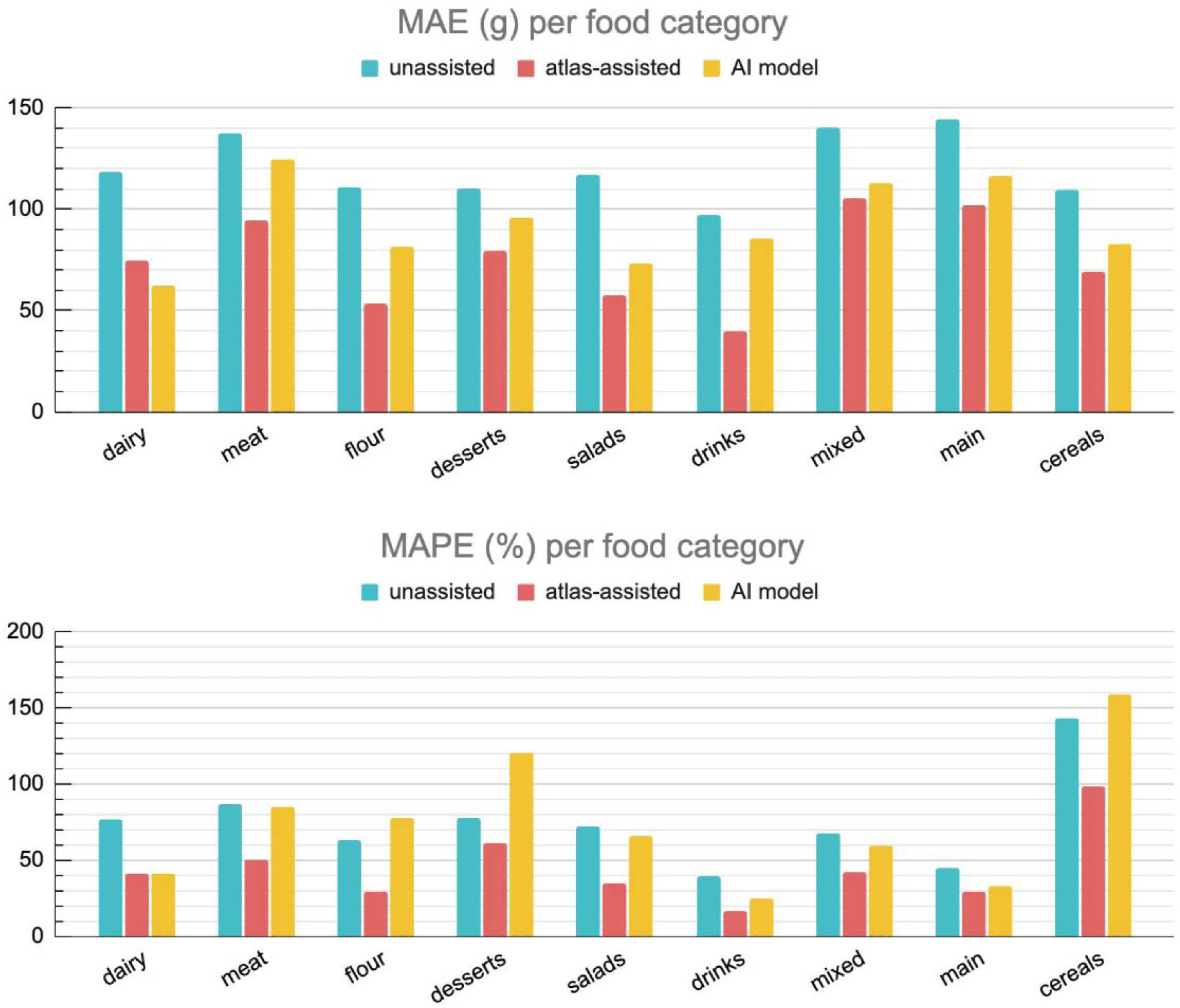
This figure shows mean absolute error (MAE) and mean absolute percentage error (MAPE) across nine food categories. Food categories included meat-based, dairy-based, flour-based, desserts, salads, mixed dishes, main dishes, drinks, and cereals.

Across all food categories except dairy, the atlas-assisted method consistently demonstrated lower MAE values than unassisted estimation, indicating improved accuracy when standardized visual references were available. The greatest reductions in MAE were observed for salads, drinks, and flour-based dishes, where visual cues likely facilitated a more accurate assessment of amorphous or variably plated items. Notably, in the salads category, atlas-assisted estimation reduced MAE by approximately 50% compared to the unassisted method.

The AI-assisted method demonstrated variable performance, generally falling between the unassisted and atlas-assisted methods in MAE across most categories. However, it exhibited particularly high MAPE in the cereals category, exceeding 150%, indicating poor proportional accuracy despite moderate absolute errors. This suggested challenges in estimating small or lightweight portions, which were especially sensitive to small errors. Conversely, the AI model performed better in drinks and main dishes, where portions were more visually distinct and consistent.

These results underscored the utility of the visual food atlas as a reliable, culturally adapted tool, especially for diverse food types. While the AI model showed potential, particularly in structured food categories, it required further refinement for improved performance in foods with high visual or portion variability, such as cereals.

### Focused Analysis: Meat and Dairy Categories

Given the centrality of meat and dairy in Central Asian diets (35), a detailed analysis was conducted for these categories. **Table 3** summarizes MAE and MAPE values by portion size and method, with visualization in **Figure 5**.

**Table 3.** MAE and MAPE for meat- and dairy-based food items.

|  |  | Unassisted |  | Atlas-assisted |  | AI Model |  |
| --- | --- | --- | --- | --- | --- | --- | --- |
|  |  | MAE (g) | MAPE (%) | MAE (g) | MAPE (%) | MAE (g) | MAPE (%) |
| **Meat-based categories** | **Overall** | 154.11 | 86.46 | 94.53 | 50.05 | 113.63 | 86.42 |
|  | **Small portion** | 93.82 | 86.31 | 52.27 | 44.05 | 96.15 | 144.55 |
|  | **Average portion** | 159.67 | 96.66 | 89.76 | 56.29 | 103.35 | 72.34 |
|  | **Large portion** | 208.84 | 76.40 | 141.56 | 49.79 | 141.39 | 42.37 |
| **Dairy-based categories** | **Overall** | 145.12 | 93.70 | 86.48 | 48.65 | 94.43 | 63.70 |
|  | **Small portion** | 113.92 | 142.31 | 60.23 | 66.62 | 103.36 | 125.05 |
|  | **Average portion** | 125.90 | 70.82 | 74.52 | 38.03 | 75.19 | 38.47 |
|  | **Large portion** | 207.05 | 81.69 | 131.85 | 47.68 | 104.75 | 27.57 |

**Figure 5:**
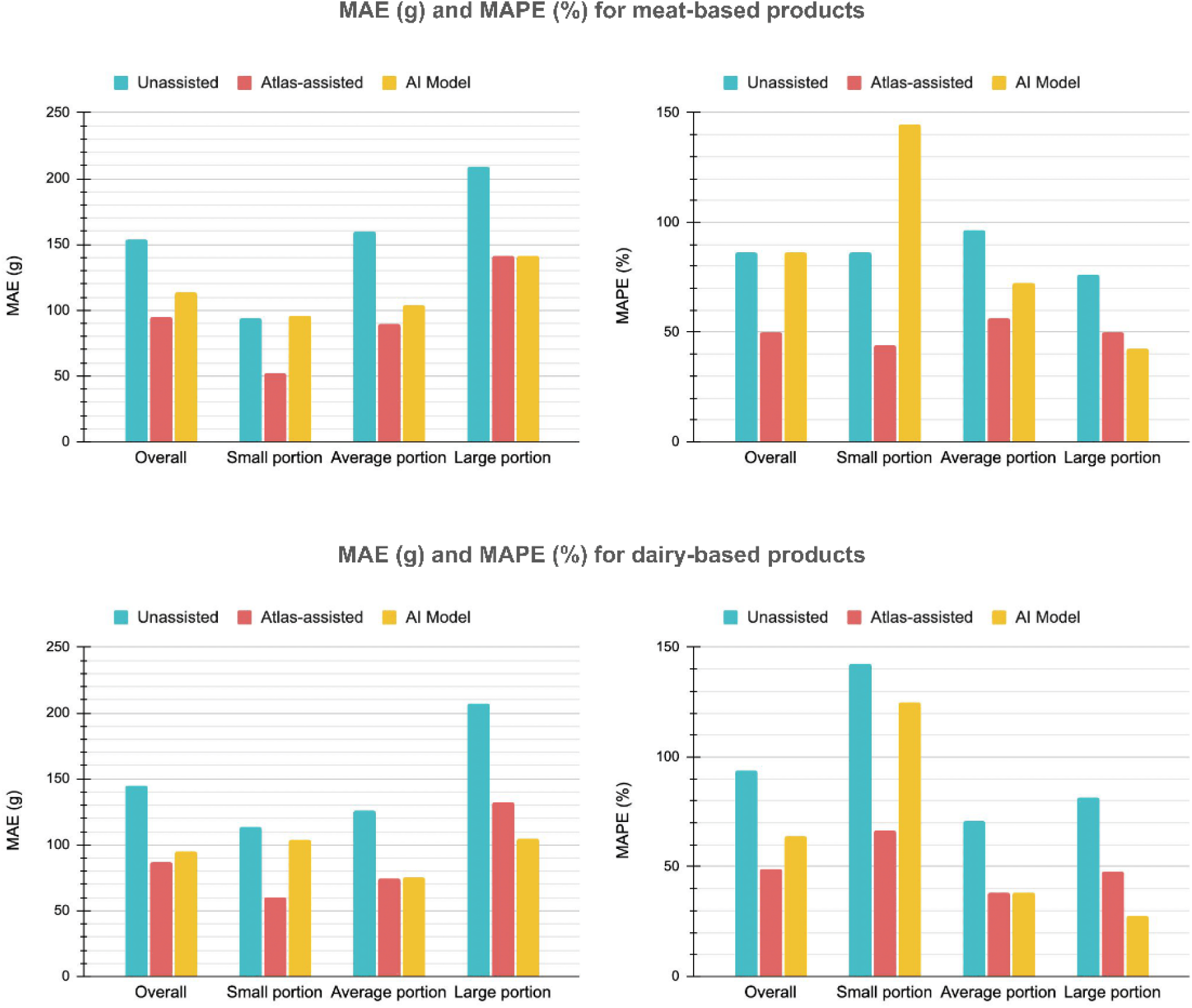
Accuracy of portion size estimation in meat- and dairy-based foods by method and portion size. Comparison of MAE and MAPE was done across unassisted, atlas-assisted, and AI model methods. Data are shown separately for small, medium, and large portions of meat-based and dairy-based items.

Across both categories and portion sizes, the atlas-assisted method consistently produced the lowest MAE and MAPE values, indicating superior accuracy over unassisted and AI-assisted estimation. In meat-based foods, the atlas-assisted approach reduced overall MAE from 154.11 g (unassisted) to 94.53 g and MAPE from 86.46% to 50.05%. This accuracy advantage was particularly pronounced in small portions, where atlas-assisted estimation reduced MAPE by nearly 50% from 86.31% to 44.05%.

The AI-assisted model demonstrated variable performance across portion sizes. For meat-based dishes, it performed comparably to atlas-assisted estimation in large portions (MAE = 141.39 g vs. 141.56 g; MAPE = 42.37% vs. 49.79%), but less accurately for small portions, where MAPE increased to 144.55%. This indicated that the AI model was more accurate in estimating larger, visually distinct portions but struggled with smaller or less visually prominent servings, leading to high proportional error.

Similar trends were observed in dairy-based foods. The atlas-assisted method again produced the lowest overall MAE (86.48 g) and MAPE (48.65%), outperforming both the unassisted method (MAE = 145.12 g; MAPE = 93.70%) and the AI model (MAE = 94.43 g; MAPE = 63.70%). Notably, the AI model showed substantial error in estimating small dairy portions, with MAPE reaching 125.05%, compared to 66.62% for the atlas-assisted method and 142.31% for unassisted estimation. In contrast, the AI model achieved the lowest MAPE among all methods for large dairy portions (27.57%), suggesting its relative strength in predicting larger, visually consistent quantities.

Overall, these findings reinforce the value of culturally adapted visual food atlases, which provide the most stable and accurate estimates across food types and portion sizes. The AI model showed promise for large portions but remains less reliable for smaller, heterogeneous servings. This limitation may be addressed with further training on small-portion food images and improved visual segmentation algorithms.

## Discussion

This study aimed to evaluate the accuracy of food portion size estimations using three different methods: unassisted human judgment, atlas-assisted estimation, and AI-assisted estimation, across a diverse set of food categories and portion sizes. The findings demonstrated that the atlas-assisted method consistently outperformed both unassisted and AI-based methods, particularly in reducing estimation errors for small and average portion sizes. This aligns with previous research indicating the value of visual references in improving estimation accuracy (19,23,37).

From an error-metric perspective, unassisted estimations consistently yielded the highest MAE and MAPE across most food categories, confirming the inherent challenges of accurate portion estimation without reference aids. The highest errors were observed in larger portions, where perceptual biases and anchoring effects are likely to contribute to overestimation. This finding aligns with the literature that suggests larger portion estimates are often skewed by visual cues and scale perception (37).

In contrast, the atlas-assisted method significantly improved portion estimation accuracy, especially for small and standard portion sizes. Its consistent performance across diverse food items, including amorphous and culturally unfamiliar dishes, underscored the utility of structured, visual aids in minimizing estimation bias in populations unfamiliar with standardized portioning. These findings further confirmed the effectiveness of culturally adapted visual tools, especially in regions where traditional eating practices involve communal sharing of food from large platters. This tool could contribute to more reliable dietary data, which is critical for informing public health interventions aimed at reducing the prevalence of non-communicable diseases in Central Asia.

While the AI model showed competitive performance for estimating large portions (particularly in dairy-based dishes), it demonstrated variable results across food categories and portion sizes. In particular, the AI model struggled with estimating small portions, especially for meat-based foods, where MAE and MAPE exceeded 140%. This discrepancy suggested that the model’s performance was hindered by smaller, more complex food types, where visual cues alone may not have been sufficient for accurate estimation. These limitations are consistent with previous findings that AI systems require large and diverse datasets for training to handle complex food structures (38). Despite these challenges, the AI model showed promise for larger, more visually distinct portions, such as those in dairy-based foods, where it achieved the lowest MAPE (27.57%) for large portions. This indicated the potential for AI to be used more effectively for standardized food types, though further refinement is needed for smaller and more complex foods.

The analysis by food category revealed further nuances. Atlas-assisted estimation consistently outperformed unassisted and AI methods across all categories, with particularly large improvements in salads, drinks, and flour-based dishes. In contrast, the AI model performed well in drinks and some standard foods but exhibited significant variability in estimating desserts and cereals, where both MAE and MAPE spiked. These findings emphasized the importance of food structure and visual complexity in determining the accuracy of portion size estimation methods. As such, the method’s efficacy was not solely dependent on the tool used but also on the nature of the food being estimated.

Further analysis of the meat-based and dairy-based food groups highlighted similar trends, with the atlas-assisted method again yielding the lowest errors in both groups. In contrast, the AI model showed competitive performance, but with substantial variability, especially in meat and dairy products with complex textures. These results are particularly relevant to Central Asian dietary patterns, where meat and dairy are central to daily diets and often involve complex textures and traditional cooking methods (35). The AI model’s higher error rates in small portions suggest that further training on small-portion food images and more contextual data (such as density, texture, and preparation methods) may be required to improve accuracy in this context.

This study had several limitations. The sample consisted predominantly of younger adults (18-35 years old) from urban Astana, Kazakhstan, which might have limited the applicability of the findings to older populations or rural settings with different dietary habits. The food items used in this study were selected to represent a range of textures and food types, but the limited diversity (51 foods, 8 beverages) might not have fully captured the breadth of Central Asian culinary practices. Additionally, while the AI model demonstrated superior accuracy overall, it struggled with small portion sizes and certain complex food types, such as dense or irregularly plated foods. This suggested that reliance on visual data alone, without supplementary information on density, texture, and cooking methods, limited the AI model’s ability to account for portion variations.

This study presented a promising foundation for the development of more region-specific dietary assessment tools. To enhance the accuracy and applicability of visual food atlases and AI models, future research should expand the dataset to include more Central Asian foods, particularly small portions, liquid-based dishes, and traditional foods. Training the AI model with these types of foods will likely improve its adaptability to a broader range of food types, reducing errors in estimating portions.

Moreover, as smartphone usage increases in urban areas, there is a growing opportunity for mobile-based dietary tracking tools that combine AI-based image recognition with real-time nutrition tracking apps. This would allow users to track their food intake simply by taking photographs, making dietary assessments more objective and time-efficient. AI-based dietary tools could thus help overcome recall bias and enhance the quality of dietary data collection, especially in real-world public health settings.

Finally, further research is needed to test AI-based dietary tools in different cultural settings. Cross-cultural comparisons between regions with different communal dining practices, such as South Asia, Sub-Saharan Africa, and the Middle East, could identify the strengths and limitations of AI-based methods and visual food atlases in global health assessments.

## Acknowledgements

BN, ZO, and MYC designed research; BN, ZO, and MYC conducted research; BN, ZO, and AS analyzed data; BN, ZO, and MYC wrote the paper; BN, ZO, MYC, and HAV reviewed and edited the paper; AS and HAV contributed visualization (other); HAV and MYC contributed resources (other); MYC supervised and administered the project (other). All authors read and approved the final manuscript and had primary responsibility for final content.

## Declaration of Generative AI and AI-assisted technologies in the writing process

During the preparation of this work, the authors used ChatGPT in order to improve the readability of some sentences. After using this tool/service, the authors reviewed and edited the content as needed and take full responsibility for the content of the publication.

## Data Availability Statement

All data produced in the present study are available upon reasonable request to the authors

## Abbreviations

AI: artificial intelligence
AR: augmented reality
CAFD: Central Asian Food Dataset
CAFSD: Central Asian Food Scenes Dataset
Fpb: Food Portion Benchmark
Ivan: Immersive Virtual Alimentation And Nutrition
iVR: immersive virtual reality
MAE: mean absolute error
MAPE: mean absolute percentage error
NCD: non-communicable disease
NUSOM IREC: Nazarbayev University School of Medicine Institutional Research Ethics Committee
SD: standard deviation

